# Are primary care virtual visits associated with higher emergency department use? A cross-sectional analysis from Ontario, Canada

**DOI:** 10.1101/2022.09.08.22278709

**Authors:** Tara Kiran, Michael E. Green, Rachel Strauss, C. Fangyun Wu, Maryam Daneshvarfard, Alexander Kopp, Lauren Lapointe-Shaw, Lidija Latifovic, Eliot Frymire, Richard H. Glazier

**Author notes:** **Corresponding author** Tara Kiran, MD, MSc, MAP Centre for Urban Health Solutions, St. Michael’s Hospital, Unity Health Toronto, 30 Bond St., Toronto, ON Canada M5B 1W8. **Data sharing:** The data sets from this study are held securely in coded form at ICES. Data-sharing agreements prohibit ICES from making the data sets publicly available, but access may be granted to those who meet pre-specified criteria for confidential access, available at www.ices.on.ca/DAS. The complete data set creation plan, and underlying analytic code are available from the authors upon request, understanding that the programs may rely upon coding templates or macros unique to ICES. **Author contributions** TK and RG conceived of the study. All authors helped design the study. FW, AK, RS and LL conducted the analysis. All authors helped interpret the data. TK and RS drafted the manuscript and all authors critically reviewed it. All authors read and approved the final manuscript.

## Abstract

**Importance:** The COVID-19 pandemic has resulted in increased use of virtual care, however, few studies have looked at the association between virtual primary care visits and other healthcare use.

**Objective:** To determine whether there was an association between a high proportion of virtual visits in primary care and more emergency department visits

**Design:** A cross-sectional study, using routinely collected data

**Setting:** Ontario, Canada

**Participants:** Ontario residents alive on March 31^st^ 2021 and family physicians with at least 1 visit claim between February and October 2021.

**Exposure:** Family physicians stratified by the percentage of total visits that were virtual (phone or video) between February and October 2021

**Main outcome(s) and measure(s):** We calculated the emergency department visit rate for each stratum of family physician virtual care use. We used multivariable logistic regression models to understand the relative rate of patient emergency department use after stratifying for rurality and adjusting first for patient characteristics and then the 2019 emergency department visit rate.

**Results:** We analyzed data for 15,155 family physicians and 12,951,063 Ontarians attached to these physicians. The mean number of emergency department visits was highest among patients whose physicians provided only in-person care (470.3 ± 1918.8 per 1,000) and was lowest among physicians who provided >80 to <100% care virtually (242.0 ± 800.3 per 1,000). After adjustment for patient characteristics patients seen by physicians with >20% of visits delivered virtually had lower rates of emergency department visits compared to patients of physicians who provided >0%-20% virtually (e.g. >80 to <100% vs >0%-20% virtual visits in Big Cities, Relative Rate (RR) 0.80 [95%CI 0.76-0.83]). This trend held across all rurality strata and after adjustment for 2019 emergency department visit rates. In urban areas, there was a gradient whereby physicians providing the highest level of virtual care had the lowest emergency department visit rates.

**Conclusions and Relevance:** Physicians who provided a high proportion of care virtually did not have higher emergency department visits than those who provided the lowest levels of virtual care. Our findings refute hypotheses that emergency department use is being driven by family physicians providing more care virtually.

**Key points:** *Question:* Do family physicians who provide more care virtually have higher emergency department visit rates among their patient panel?

*Findings:* In this cross-sectional study from Ontario, Canada, we examined data from February to October 2021 for 12,951,063 patients attached to 15,155 family doctors and found that physicians who provided a high proportion of virtual care did not have higher emergency department visits than those who provided the lowest levels of virtual care. This finding remained true after adjusting for patient characteristics.

*Meaning:* Our findings refute hypotheses that emergency department use is being driven by family physicians providing more care virtually.

## Introduction

The COVID-19 pandemic resulted in a seismic shift in how primary care was delivered. Shortly after a global pandemic was declared in March 2020, the amount of care delivered virtually—by phone or video—increased 56-fold, comprising over 70% of all primary care visits.^1^ More than two years into the pandemic, virtual care continues to comprise a substantial portion of visits.^2^ Research has found that most patients feel comfortable with virtual care, like its convenience, and want it to continue.^3-6^ However, there are concerns about the impact of virtual primary care visits on care quality including on patient safety,^7^ equity in access,^8,9^ effective chronic condition management,^10^ and healthcare utilization.^11^ Effects have been hard to untangle due to the simultaneous impact of the pandemic on care.

The proportion of primary care delivered virtually varies by jurisdiction and practice and is influenced by factors such as reimbursement and overhead, patient and provider access to technology and infrastructure, provider access to personal protective equipment, provider health concerns, and patient and provider preference.^12-17^ There have been concerns that in some practices, the proportion of virtual visits is too high resulting in an increase in other health system use. However, few studies have looked at the association between virtual primary care visits and other healthcare use. One such study found high practice telehealth use was associated with a small increase in emergency room visits and hospitalizations for ambulatory care sensitive conditions compared with low practice telehealth use.^11^

In Ontario, Canada, provincial authorities had initially directed family physicians to adopt a virtual-first approach to seeing patients as a safety precaution when COVID-19 case counts were high. This approach meant physicians were to perform the initial assessment remotely and then bring patients into the office for an in-person visit only if warranted. However, by fall 2021, authorities were concerned that many family physicians were doing too much virtual care and that this was making it difficult for patients to access in-person care, resulting in increased emergency department use. In October 2021, the provincial medical officer of health and the regulatory college issued a joint statement encouraging more use of in-person care.^18^ It is unclear, however, whether high use of virtual care was driving perceived increases in emergency department visit volumes.

We undertook a cross-sectional study using routinely collected data to understand whether there was an association, ecologically and at a practice-level, between a high proportion of virtual visits in primary care and more emergency department visits.

## Methods

### Context and Setting

Ontario is Canada’s largest province with a population of 14.8 million in 2021.^19^ Physician and hospital visits, including emergency department care, are fully insured and free at the point-of-care for all permanent residents through the Ontario Health Insurance Plan (OHIP). Just over 80% of the population is formally enrolled to a family physician practicing in a Patient Enrolment Model where, depending on the model of care, between 15% to 70% of payment is via capitation (adjusted for patient age and sex) with some fee-for-service and incentive payments.^20^ The remaining population either are unattached or see a family physician practicing fee-for-service. Approximately one quarter of the population lives in rural communities with fewer than 30,000 people, where health services are structured differently.^21^ Rural emergency departments are staffed by family physicians and although emergency department volumes are relatively low, visit rates are much higher than in urban areas, due in part to the limited availability of other after-hours care.^22^ Temporary virtual care billing codes were introduced for phone and video visits in Ontario on March 14, 2020 and, during the study time period, paid the same amount as an equivalent in-person visit. Initially, the same billing code was used for phone and video.

### Study Design

We examined primary care and emergency department visit trends between January 2019 and October 2021. We then conducted a cross-sectional, population-based analysis using linked health administrative data to examine the association between the proportion of care delivered virtually at the primary care physician-level and emergency department visit rates between February and October 2021 in Ontario, Canada.

Datasets were linked using unique encoded identifiers and analyzed at ICES. ICES is an independent, non-profit research institute whose legal status under Ontario’s health information privacy law allows it to collect and analyze health care and demographic data, without consent, for health system evaluation and improvement.

### Study Population

We examined visit trends for all residents in Ontario. For the cross-sectional analysis, we included family physicians with at least 1 home, office, or virtual visit claim between February and October 2021. We included permanent residents of Ontario who were alive as of March 31^st^ 2021. We assigned Ontario residents to a family physician using enrolment tables from the Ontario Ministry of Health; for those not enrolled, we assigned them to the family physician who billed the maximum value of 30 common primary care fee codes billed within the previous 2 years (eExhibit 1). We excluded residents who we could not assign to a family physician or whose family physician did not have any visit claims during the study period.

We categorized physicians as practicing traditional fee-for-service or in one of the following three types of Patient Enrolment Models: *Enhanced fee-for-service* (Comprehensive Care Model, Family Health Group) where fee-for-service comprises approximately 80% of income; *Non-team capitation* (Family Health Network, Family Health Organization) where capitation comprises approximately 70% of income; and *Team-based capitation* (Family Health Team) where capitation comprises approximately 70% of income and physicians also have access to an interprofessional team funded through the Ministry of Health.

### Analysis

#### Trends in primary care and emergency department visits

We used physician billing claims to count primary care visits weekly by type: office, home, virtual and total. We used the National Ambulatory Care Reporting System to assess emergency department visits weekly (total and stratified by triage level). For context, we included data on COVID-19 case counts in Ontario using publicly available COVID-19 surveillance data from Public Health Ontario.^23^

#### Cross-sectional description of physicians by virtual care use

For each family physician, we calculated the total volume of visits that were in-person (office and home) and virtual (phone and video) between February and October 2021. We then calculated the percent of all visits that were delivered virtually during that period and stratified physicians into the following groups *a priori*: 0% virtual (100% in-person), >0-20%, >20-40%, >40-60%, >60-80%, >80-<100%, and 100% virtual.

We described patient and physician characteristics as of March 31^st^, 2021 within each stratum of virtual care use. We assessed physician characteristics using the provider databases. We assessed patient age, sex, and postal code using the provincial registry of all individuals eligible for OHIP coverage. We used postal code and 2016 Canadian census data to derive neighbourhood-level income quintile. We assessed rurality using patient and physician postal code and the Rurality Index of Ontario (RIO) score, where areas with a score of 0 are considered big cities, 1-9 are small cities, 10-39 are small towns, and 40 or more are rural areas.^24^ Patient postal code and the Ontario Marginalization Index^25,26^ were used to assess neighbourhood-level material deprivation and ethnic diversity. We assessed whether patients registered for OHIP within the last ten years, a proxy for recent immigration. Finally, we used the Johns Hopkins Adjusted Clinical Group method to assess patient co-morbidity and morbidity (ACG® System Version 10).^27^ We used the Johns Hopkins ACG® System Aggregated Diagnosis Groups to categorize comorbidity as 0 (no comorbidity), 1-4 (low comorbidity), 5-9 (moderate comorbidity) and 10+ (high comorbidity). Morbidity was assessed using corresponding Resource Utilization Bands (RUBs) categorized as 0-1 (non-user/healthy user), 2 (low morbidity), 3 (moderate morbidity) and 4+ (high morbidity).^28^

#### Variation in virtual care use

For physicians in a Patient Enrolment Model, we examined the variation in virtual care use between physicians in the same practice group and between physicians in different practice groups, stratified by the type of Patient Enrolment Model. For this analysis, the percentage of total visits that were virtual was modeled as a continuous variable for each physician. To understand how much of the total variance in virtual visits was attributable to physician group and practice type, we calculated an intraclass correlation coefficient from a three-level (patient, physician and group), intercept-only mixed linear model with virtual visit rate as the outcome.

#### Cross sectional association between virtual care and other health care use

We examined healthcare use of patients attached to family physicians within stratum of virtual care use. The primary outcome was the rate of emergency department visits per 1,000 patients. We compared this across levels of family physician virtual care use between February and October 2021. For each stratum, we assessed the number of emergency department visits for every patient of physicians in the stratum and calculated the average number per patient. We also assessed the following contextual information: i) the percent of patients with a primary care visit, ii) the average number of primary care visits, and iii) the percentage of visits to the usual family physician between February and October 2021. Usual family physician was the physician a patient saw the most over the two years prior to March 31, 2021. Secondary outcomes included the percent of patients with an ambulatory care sensitive condition-related hospital admission, derived from the Canadian Institute for Health Information’s Discharge Abstract Database (eExhibit 2), and specialist visits, ascertained from physician billings. We examined bivariate associations overall and separately for each rurality stratum.

Next, we wanted to understand whether the associations observed in 2021 reflected historical associations that pre-dated the pandemic and widespread adoption of virtual care. To do so, we examined the absolute and relative differences in the emergency department visit rate between February and October 2021 and the corresponding period (February to October) in 2019 for each stratum of family physician virtual care use.

Finally, we aimed to understand the relationship between physician virtual care use and patient emergency department visits after adjustment for potential confounders. We constructed two multivariable logistic regression models with physician-level virtual care proportion as a 5-level categorical exposure and volume of patient emergency department visits as the outcome. In Model 1, we adjusted for patient age, sex, neighbourhood income quintile, recent registration, co-morbidity, and healthcare use, and stratified by rurality. Model 2 included the same covariates and further adjusted for the emergency department visit rate in 2019, to account for potential pre-existing patterns. All analyses were performed in SAS EG 7.1.

#### Sensitivity Analysis

We performed sub-group analyses where we used a previously-published algorithm^29^ to restrict our cohort to comprehensive primary care physicians, active as of March 31, 2019, the most recent year where the data for the algorithm was available.

### Ethics Approval

The use of the data in this project is authorized under section 45 of Ontario’s Personal Health Information Protection Act (PHIPA) and does not require review by a Research Ethics Board.

## Results

### Primary Care and Emergency Department Visits Over Time

Total primary care visits dropped at the onset of the pandemic but returned to average pre-pandemic levels by fall of 2020 (Figure 1a). The proportion of virtual primary care visits peaked in the first two weeks of the pandemic at 82% but was 49% by October 2021. Emergency department visits dropped at the start of the pandemic and remained lower than 2019 volumes throughout the study period (Figure 1b). Trends in ED visit rates were similar for Canadian Triage and Acuity Scale (CTAS) levels 2,3, and 4 (Appendix, eExhibit 3) with dips visually corresponding to increases in COVID numbers (Figure 1c). Periods with a higher proportion of virtual primary care visits visually related to higher numbers of COVID cases and lower ED visit rates (Figures 1 c and d).

**Figure 1.**
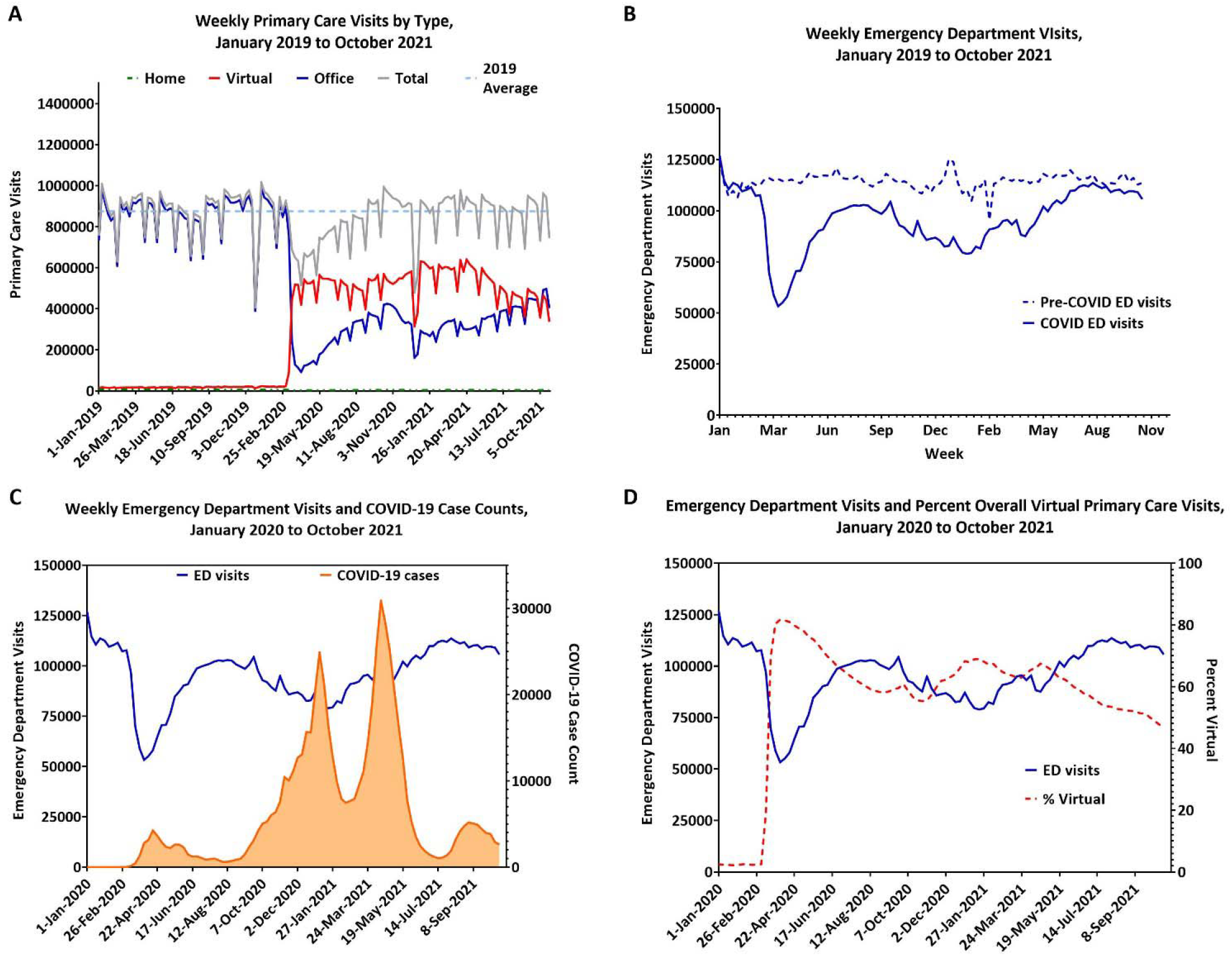
Primary Care Visits, Emergency Department Visits, and Percent of Total Primary Care Visits Delivered Virtually, January 2019 to October 2021. *(A): Weekly primary care visits, by type of visit, over time. (B): Comparison of weekly emergency department visits during the Pre-COVID-19 period (January 2019 – October 2019) and the COVID-19 period (January 2020 to October 2021). (C): Comparison of weekly emergency department visits and COVID-19 case counts between January 2020 and October 2021. (D): Comparison of weekly emergency department visits and percent of primary care visits that were virtual between January 2020 and October 2021.*

### Physician and Patient Characteristics

We analyzed data for 15,155 family physicians who had at least one office, home, or virtual claim between February and October 2021 and 12,951,063 Ontarians who were attached to these physicians (Table 1). The majority of physicians provided between 40-80% of care virtually; only 400 physicians provided 100% of care virtually while 2,691 provided between >80 to <100% of care virtually. A higher proportion of the physicians who provided >80% virtual care were 65 years or older, female, and practicing in big cities. Most physicians who provided 100% of care virtually worked traditional fee-for-service and had a panel size of less than 100; more of their patients were recent registrants and lived in a neighbourhood in the lowest income quintile and a neighbourhood that was more ethnically diverse (Appendix, eExhibit 4). Patient co-morbidity and morbidity were similar across strata.

**Table 1.** Physician and Practice Characteristics by the Percent of Care Provided Virtually, between February 1 and October 31, 2021, in Ontario, Canada.

| Physician % Virtual Care | 0% | >0-20% | >20-40% | >40-60% | >60-80% | >80-<100% | 100% | Total |
| --- | --- | --- | --- | --- | --- | --- | --- | --- |
| **No. Physicians** | N=1,395 | N=1,447 | N=1,861 | N=3,099 | N=4,262 | N=2,691 | N=400 | N=15,155 |
| **No. Patients** | N=82,689 | N=771,808 | N=1,633,324 | N=3,229,223 | N=4,657,341 | N=2,511,964 | N=64,714 | N=12,951,063 |
| **Physician Age** |  |  |  |  |  |  |  |  |
| ≤44 | 629 (45.1) | 529 (36.6) | 792 (42.6) | 1,412 (45.6) | 1,868 (43.8) | 984 (36.6) | 149 (37.3) | 6,363 (42.0) |
| 45-64 | 550 (39.4) | 646 (44.6) | 788 (42.3) | 1,323 (42.7) | 1,890 (44.3) | 1,160 (43.1) | 133 (33.3) | 6,490 (42.8) |
| 65-74 | 148 (10.6) | 202 (14.0) | 221 (11.9) | 300 (9.7) | 421 (9.9) | 427 (15.9) | 81 (20.3) | 1,800 (11.9) |
| ≥75 | 50 (3.6) | 70 (4.8) | 60 (3.2) | 64 (2.1) | 83 (1.9) | 120 (4.5) | 37 (9.3) | 484 (3.2) |
| **Physician Sex** |  |  |  |  |  |  |  |  |
| Female | 526 (37.7) | 499 (34.5) | 778 (41.8) | 1,590 (51.3) | 2,393 (56.1) | 1,355 (50.4) | 214 (53.5) | 7,355 (48.5) |
| Male | 869 (62.3) | 948 (65.5) | 1,083 (58.2) | 1,509 (48.7) | 1,869 (43.9) | 1,336 (49.6) | 186 (46.5) | 7,800 (51.5) |
| **Primary Care Model** |  |  |  |  |  |  |  |  |
| PEM: Non-team capitation | 14 (1.0) | 148 (10.2) | 354 (19.0) | 788 (25.4) | 1,267 (29.7) | 570 (21.2) | 11 (2.8) | 3,152 (20.8) |
| PEM: Enhanced fee-for-service | 31 (2.2) | 194 (13.4) | 332 (17.8) | 492 (15.9) | 923 (21.7) | 777 (28.9) | 32 (8.0) | 2,781 (18.4) |
| PEM: Team-based capitation | 31 (2.2) | 102 (7.0) | 377 (20.3) | 980 (31.6) | 1,172 (27.5) | 360 (13.4) | 12 (3.0) | 3,034 (20.0) |
| Traditional fee-for-service | 816 (58.5) | 827 (57.2) | 665 (35.7) | 674 (21.7) | 741 (17.4) | 904 (33.6) | 285 (71.3) | 4,912 (32.4) |
| Missing | 503 (36.1) | 176 (12.2) | 133 (7.1) | 165 (5.3) | 159 (3.7) | 80 (3.0) | 60 (15.0) | 1,276 (8.4) |
| **Location of Practice** |  |  |  |  |  |  |  |  |
| Big cities | 567 (40.6) | 623 (43.1) | 736 (39.5) | 1,266 (40.9) | 2,217 (52.0) | 1,565 (58.2) | 251 (62.8) | 7,225 (47.7) |
| Small cities | 297 (21.3) | 381 (26.3) | 458 (24.6) | 816 (26.3) | 1,194 (28.0) | 778 (28.9) | 86 (21.5) | 4,010 (26.5) |
| Small towns | 303 (21.7) | 258 (17.8) | 373 (20.0) | 629 (20.3) | 523 (12.3) | 228 (8.5) | 31 (7.8) | 2,345 (15.5) |
| Rural | 210 (15.1) | 185 (12.8) | 294 (15.8) | 388 (12.5) | 328 (7.7) | 120 (4.5) | 32 (8.0) | 1,557 (10.3) |
| Missing | 18 (1.3) | 0 (0.0) | 0 (0.0) | 0 (0.0) | 0 (0.0) | 0 (0.0) | 0 (0.0) | 18 (0.1) |
| **Patient Panel Size** |  |  |  |  |  |  |  |  |
| <100 | 773 (55.4) | 592 (40.9) | 450 (24.2) | 469 (15.1) | 479 (11.2) | 584 (21.7) | 242 (60.5) | 3,589 (23.7) |
| 100-499 | 72 (5.2) | 230 (15.9) | 235 (12.6) | 278 (9.0) | 363 (8.5) | 350 (13.0) | 54 (13.5) | 1,582 (10.4) |
| 500-999 | 18 (1.3) | 127 (8.8) | 261 (14.0) | 608 (19.6) | 897 (21.0) | 493 (18.3) | 25 (6.3) | 2,429 (16.0) |
| 1000-1499 | 21 (1.5) | 120 (8.3) | 356 (19.1) | 722 (23.3) | 1,214 (28.5) | 546 (20.3) | 9 (2.3) | 2,988 (19.7) |
| 1500-1999 | ≤5 | 85 (5.9) | 225 (12.1) | 489 (15.8) | 697 (16.4) | 348 (12.9) | ≤5 | 1,855 (12.2) |
| ≥2000 | ≤5 | 117 (8.1) | 201 (10.8) | 368 (11.9) | 453 (10.6) | 290 (10.8) | ≤5 | 1,436 (9.5) |
| Missing | 503 (36.1) | 176 (12.2) | 133 (7.1) | 165 (5.3) | 159 (3.7) | 80 (3.0) | 60 (15.0) | 1,276 (8.4) |

Note: Ontario residents who were not attached to a primary care physician or whose physician did not have claims during the study period were excluded.
PEM= Patient Enrolment Model

### Variation in the proportion of virtual care

Among physicians practicing in a patient enrolment model, we found substantial variation in the proportion of virtual care provided both between and within groups of physicians (Figure 2). The variation was not explained by practice model (Interclass Correlation Coefficient for practice model = 0.5%); physician group accounted for almost one-third of the observed variation between all physicians (Interclass Correlation Coefficient for physician group = 30.5%).

**Figure 2.**
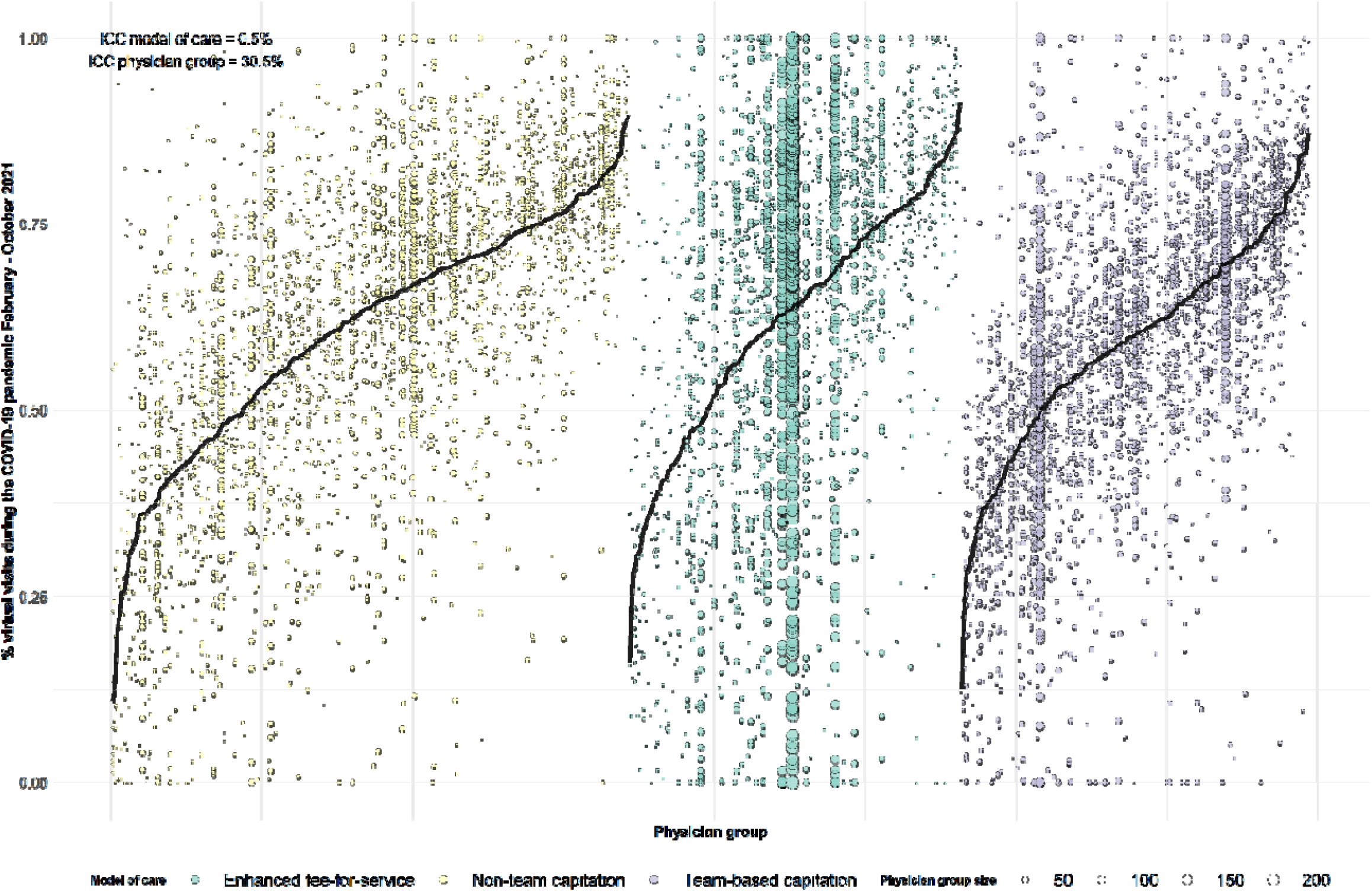
Variation in the Percent of Virtual Visits between Physicians in the Same Group and between Physicians in Different Groups, Stratified by Type of Patient Enrolment Model, February and October 2021 *The black line represents the mean ratio for the practice group. Each group can have 3 or more mos. Each dot represents a physician. Physicians within the same practice group are represented on the same vertical line. We calculated an intraclass correlation coefficient from a three-level, intercept-only mixed linear model to understand how much of the total variance in virtual visits was attributable to physician group and practice type. We found the variation was not explained by model of care (ICC: 0.5%), whereas a high proportion of variation was explained by specific practice group the physician belonged to (ICC:30.5%).*

### Health Service Utilization

Compared to patients whose physicians provided 40-60% of care virtually, patients whose physicians provided >80% of care virtually had a higher mean number of primary care visits per patient and lower continuity; they did not have substantially different referral rates to specialists or rates of hospital admission for ambulatory sensitive conditions (Table 2).

**Table 2.** Patient Health Service Utilization Stratified by the Rostered Physician’s Percent of Care Provided Virtually During The Eight-Month Period Between February 1 and October 2021

| Physician % Virtual Care | 0% | >0-20% | >20-40% | >40-60% | >60-80% | >80-<100% | 100% | Total |
| --- | --- | --- | --- | --- | --- | --- | --- | --- |
| **No. Patients** | N=82,689 | N=771,808 | N=1,633,324 | N=3,229,223 | N=4,657,341 | N=2,511,964 | N=64,714 | N=12,951,063 |
| **Primary Care Services** |  |  |  |  |  |  |  |  |
| Any primary care visit, n (%) | 46,285 (56.0) | 476,755 (61.8) | 1,011,337 (61.9) | 2,015,294 (62.4) | 2,998,836 (64.4) | 1,652,188 (65.8) | 40,898 (63.2) | 8,241,593 (63.6) |
| Number of primary care visits per patient, mean ± SD | 2.5 ± 4.9 | 2.5 ± 4.2 | 2.4 ± 3.9 | 2.3 ± 3.7 | 2.6 ± 3.9 | 2.9 ± 4.3 | 3.1 ± 5.34 | 2.5 ± 3.9 |
| Percent of visits with most responsible family physician*, mean ± SD | 47.9 ± 44.8 | 66.09 ± 40.3 | 70.0 ± 38.2 | 71.7 ± 36.7 | 71.5 ± 36.8 | 67.9 ± 38.9 | 39.5 ± 42.5 | 70.0 ± 37.8 |
| **Other Health Care Use** |  |  |  |  |  |  |  |  |
| Any specialist visits, n (%) | 31,481 (38.1) | 290,308 (37.6) | 621,311 (38.0) | 1,251,610 (38.8) | 1,855,914 (39.8) | 1,012,442 (40.3) | 25,745 (39.8) | 5,088,811 (39.3) |
| Ambulatory care-sensitive condition visits, n (%) | 324 (0.4) | 2,085 (0.3) | 4,104 (0.3) | 7,761 (0.2) | 9,608 (0.2) | 4,699 (0.2) | 114 (0.2) | 28,695 (0.2) |
| **Emergency Department Utilization, February to October 2019/2021** |  |  |  |  |  |  |  |  |
| Total visits per 1,000, 2021, mean ± SD† | 470.3 ± 1918.8 | 309.8 ± 1087.0 | 295.3 ± 1036.1 | 283.5 ± 919.7 | 253.4 ± 825.4 | 242.0 ± 800.3 | 287.1 ± 1048.9 | 268.9 ± 903.2 |
| Total visits per 1,000, 2019, mean ± SD† | 468.3 ± 1850.5 | 360.6 ± 1100.1 | 346.5 ± 996.9 | 329.0 ± 985.8 | 290.1 ± 934.2 | 272.0 ± 843.2 | 294.0 ± 891.4 | 309.6 ± 965.4 |
| Absolute difference 2021 vs 2019 (95% CI) | 2.0 (-14.1, 18.1) | -50.7 (-54.2, -47.3) | -51.3 (-53.6, -49.0) | -45.5 (-47.0, -44.0) | -36.7 (-37.9, -35.5) | -30.0 (-31.4, -28.5) | -6.9 (-16.8, 3.0) | -40.7 (-41.44, -40.0) |
| Percent change 2021 vs 2019 (95% CI) | 0.4 (-3.0, 3.9) | -14.1 (-15.0, -13.2) | -14.8 (-15.4, -14.2) | -13.8 (-14.3, -13.4) | -12.7 (-13.0, -12.3) | -11.0 (-11.5, -10.5) | -2.4 (-5.8, 1.1) | -13.2 (-13.4, -12.9) |

\*Among patients with 2 or more visits between February and October 2021; †This includes only patients whose attached physicians had claims in both 2021 and 2019.
Abbreviations: SD, standard deviation; CI, confidence interval

The mean number of emergency department visits was highest among patients whose physicians provided only in-person care (470.3 ± 1918.8 per 1,000) and was lowest in the >80-<100% virtual care group (242.0 ± 800.3 per 1,000) (Table 2). Emergency department use decreased as physician percent of virtual care increased, except among physicians who provided 100% of care virtually where ED rates were 287.1 ± 1048.9 per 1000. Emergency department visits in 2019, prior to the COVID-19 pandemic, demonstrated a very similar pattern. Between 2019 and 2021, there was an overall 13% decrease in the mean number of emergency department visits (2019: 309.6 ± 965.4, 2021: 268.9 ± 903.2). Visits decreased across all levels of physician virtual care use, apart from a slight increase among patients attached to physicians who provided 0% virtual care; the absolute and relative decrease was smallest among patients of physicians who provided 100% virtual care followed by those whose physicians provided 80-<100% virtual care. The crude association between the proportion of care delivered virtually and ED use was consistent by rural strata (eExhibit 7).

### Regression Findings

Regression modeling excluded patients of physicians providing 0 or 100% virtual care (n= 1,395 and 400, respectively). We found that after adjustment for patient characteristics, patients seen by physicians who had more than 20% visits virtually had lower rates of emergency department visits when compared to patients of physicians who provided the least percentage of virtual care (>0-20%) (Figure 3a). For example, patients who lived in big cities who provided >60-80% of care virtually had almost 20% lower rate of emergency department visits compared to those who were seen by a physician providing >0-20% of care virtually. This trend held across all region types. In both big and small cities, we observed a gradient such that patients whose corresponding physicians provided the highest level of virtual care had the lowest rates of emergency department visits. These patterns remained even after further adjusting the model for patient emergency department visit rates in 2019 (Figure 3b).

**Figure 3.**
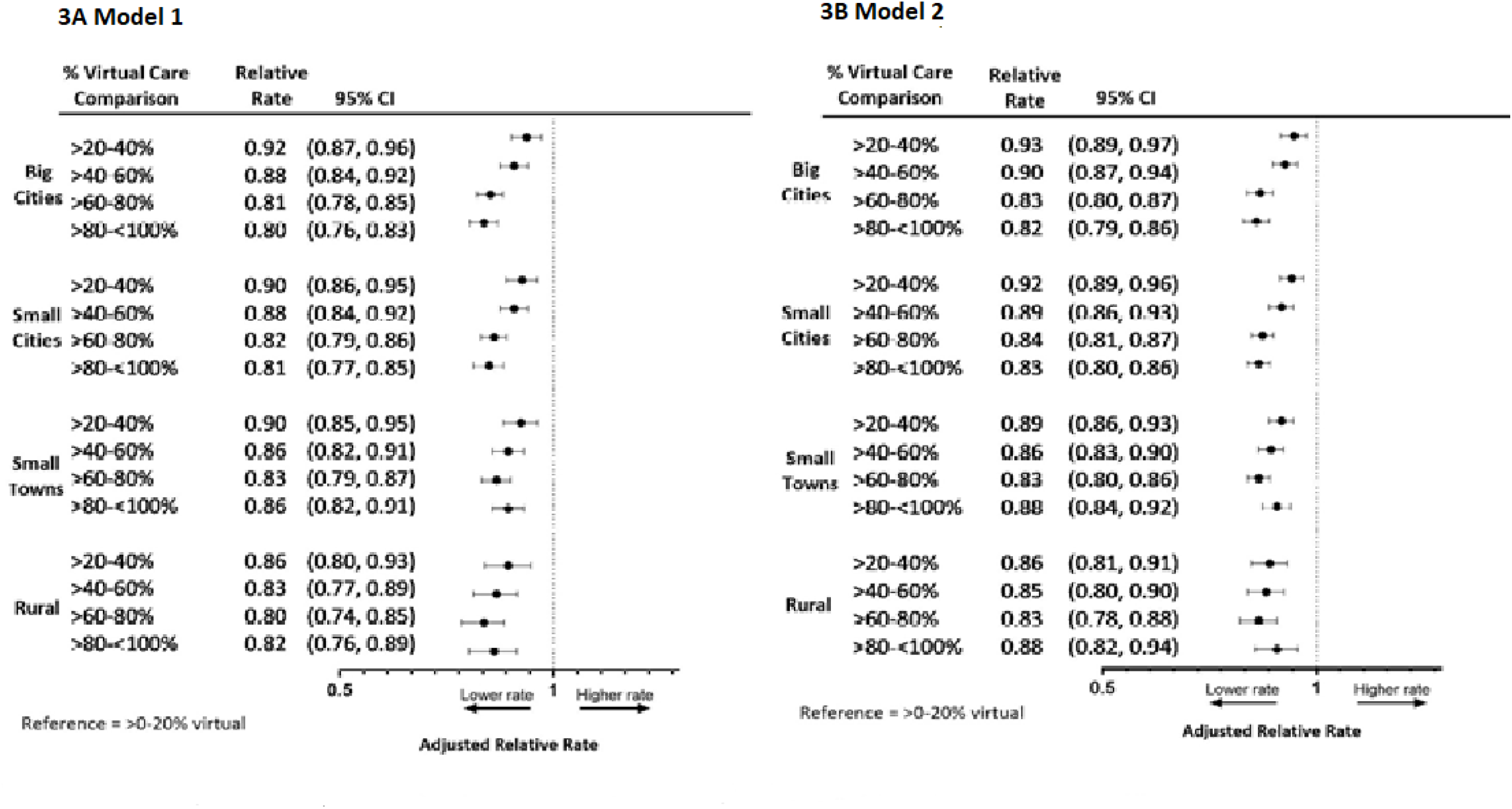
Adjusted Relative Rates of Patient Emergency Department Visits by Percent of Total Visits Delivered Virtually by Family Physicians, Stratified by Rurality. Reference = Physicians Where Virtual Care Comprises between >0 to 20% of Total Visits. *Model 1: Adjusted for patient age, sex, neighbourhood income quintile, recent registration, co-morbidity and morbidity*. *Model 2: Adjusted for patient age, sex, neighbourhood income quintile, recent registration, co-morbidity, morbidity and rate of emergency department visits in 2019*.

### Sensitivity analysis

Overall, results were consistent in the sensitivity analysis limited to comprehensive family physicians (n=9,462) (eExhibit 5 and eExhibit 6).

## Discussion

The expansion of virtual care during the pandemic opened new modes of access for patients and allowed family physicians to care for people while reducing the risk of COVID transmission. Nonetheless, some policymakers became worried that virtual care was being used inappropriately, leading to an increase in emergency department use. Our findings refute this hypothesis. First, we examined population-based trends in emergency department and primary care visits before and after the pandemic. We found emergency department visit rates were lower than in the pre-pandemic period; increases in emergency department use seemed to coincide with decreasing COVID cases and did not coincide with more virtual primary care. Second, we conducted a cross-sectional analysis of 12,951,063 million patients attached to 15,155 family doctors who practiced between February and October 2021. We found that, at the population level, physicians who provided a high proportion of virtual care did not have higher emergency department visits than those who provided the lowest levels of virtual care. This finding remained true even after adjusting for patient characteristics with differences largely following pre-pandemic patterns.

### Comparison with other studies

Prior to COVID-19, several studies suggested virtual care could reduce emergency department and other hospital use, specifically for rural populations,^30^ older populations,^31^ and following a natural disaster.^32^ Following COVID-19, a US study examining virtual care use and hospital visits for ambulatory-care sensitive conditions (ACSC) found that practices with high virtual care use had a small increase in ACSC visits compared to those with medium virtual care use, but differences disappeared when acute and chronic ACSC were evaluated separately.^11^ Other studies suggest that virtual care use during the COVID-19 pandemic was higher among patients who were sicker, suggesting it supported care continuity when COVID-19 cases were high.^1,33^ In US studies, virtual care often includes a high portion of video unlike in Ontario where most virtual care was delivered by phone.^3^

### Strengths and Limitations

We conducted a population-based analysis that included data on all family physicians in Canada’s largest province who were practicing in the study period. We examined emergency department use along with other health service use and looked back to the pre-pandemic period to understand if associations may have pre-dated the pandemic. Our findings were consistent when we limited our analysis to physicians meeting criteria for practicing comprehensive care. We examined virtual care use using new billing codes but unfortunately these do not distinguish between video and phone visits. Billings also do not capture other aspects of care such as email with patients or care provided by non-physician team members. We examined a nine-month period of the pandemic and findings may not reflect evolving practice patterns. Finally, our study is limited to administrative data and does not shed light on why there are differences in the amount of care delivered virtually, appropriateness of virtual care for specific circumstances, and whether that care meets patients’ needs.

### Implications for Policy

The pandemic resulted in the widespread adoption of virtual care, which is here to stay. In Ontario, billing for virtual care that was introduced on an emergency basis in the pandemic will become permanent in October 2022 with new billing codes that provide lower remuneration for phone appointments compared to in-person visits. However, two years into the pandemic, there continue to be news reports of emergency departments being overwhelmed and some continue to speculate that one contributing factor is patients not being able to see their physician in-person.^34^ Mixed methods research is needed to better understand patients’ experience accessing their family physicians, reasons for seeking care in the emergency department, their views on virtual care, and drivers of physician-level variation in virtual care provision. Researchers and policy-makers should be mindful of different patient subgroups where virtual care can either facilitate access (e.g. in rural areas) or be a barrier (e.g. for those with language or sensory barriers).

### Conclusion

We found that, at the population level, physicians who provided a high proportion of virtual care did not have higher emergency department visits than those who provided the lowest levels of virtual care. Further, during the first 18-months of the pandemic, emergency department visit rates were lower than pre-pandemic levels and periods where emergency department visit rates were highest did not coincide with higher rates of virtual care use. Our findings refute hypotheses that emergency department use is being driven by family physicians providing more care virtually.

## Data Availability

The data sets from this study are held securely in coded form at ICES. Data-sharing agreements prohibit ICES from making the data sets publicly available, but access may be granted to those who meet pre-specified criteria for confidential access, available at www.ices.on.ca/DAS. The complete data set creation plan, and underlying analytic code are available from the authors upon request, understanding that the programs may rely upon coding templates or macros unique to ICES.

## Appendix

**eExhibit 1.** Billing Codes Used to Determine the Usual Family Physician for Not Enrolled Patients.

| Fee code | Description |
| --- | --- |
| **Primary Care Codes** |  |
| A001 | MINOR ASSESS.-F.P./G.P. |
| A002 | Family Practice & Practice in General - Enhanced 18 month well baby visit |
| A003 | GEN. ASSESS. -F.P./G.P. |
| A007 | INTERMED.ASSESS/WELL BABY CARE-F.P./G.P./PAED. |
| A903 | GEN/FAM PRACT-PRE-DENTAL/OPER.ASSESS LIMIT 2 PER YEAR/PT |
| E075 | GERIATRIC GENERAL ASSESSMENT PREMIUM |
| G212 | D./T. PROC.-ALLERGY-HYPOSENSITIZATION INJECTION PLUS BASIC |
| G271 | D./T. PROC.-CARDIOV.-ANTICOAGULANT SUPERVISION |
| G372 | D./T. PROC.-INJECTIONS-INTRADERMAL/MUSCULAR ETC. EA. ADD. |
| G373 | D./T. PROC.-INJ. INTRADERMAL/MUSC. BASIC FEE (SHICK TEST) |
| G365 | D./T. PROC.-GYNAECOLOGY-PAPANICOLAOU SMEAR |
| G538 | D&T IMMUNIZATION-WITH VISIT, EACH INJECT. |
| G539 | Injection of unspecified agent - sole reason (first injection) |
| G590 | INFLUENZA AGENT +VISIT |
| G591 | Injection of influenza agent - sole reason |
| K005 | INDIVIDUAL CARE PER 1/2 HR |
| K013 | COUNSELLING-ONE OR MORE PEOPLE-PER 1/2HR. |
| K017 | ANNUAL HEALTH EXAM-CHILD AFT. 2ND BIRTHDAY. |
| P004 | OBS.-PRENATAL CARE-MINOR PRENATAL ASSESS.-SUBSEQ.PRENAT.VIS. |
| K130 | Periodic health visit - adolescent |
| K131 | Periodic health visit - adult aged 18 to 64 inclusive |
| K132 | Periodic health visit - adult 65 years of age and older |
| K030 | DIABETIC MANAGEMENT FEE |
| K080 | Minor assessment - Covid, Virtual |
| K081 | Intermediate assessment - Covid, Virtual |
| K082 | Primary mental health care - Covid, Virtual |
| **Paediatric Codes** |  |
| A261 | MINOR ASSESS.-PAED. |
| A268 | Paediatrics - Enhanced 18 month well baby visit |
| K267 | ANNUAL HEALTH EXAM-CHILD-AFT. 2ND BIRTHDAY PAED. |
| K269 | ANNUAL HEALTH EXAM-PAEDIATRICS-ADOLESCENT-OFFICE |

**eExhibit 2.** Ambulatory Care Sensitive Condition Related Hospital Admissions Used as Secondary Outcomes to Examine Healthcare Utilization of Patients Attached to Family Physicians within Each Stratum of Virtual Care Use.

| Most responsible diagnosis | International classification of diseases (ICD)-10-CA<sup>a</sup> code |
| --- | --- |
| Asthma | J45 |
| Diabetes | E10.0, E10.1, E10.63, E10.64, E10.9, E11.0, E11.1, E11.63, E11.64, E11.9, E13.0, E13.1, E13.63, E13.64, E13.9, E14.0, E14.1, E14.63, E14.64, E14.9 |
| Congestive heart failure | I50, J81 |
| Chronic obstructive pulmonary disease | J41, J42, J43, J44, J47, with CCI<sup>b</sup> exclusion 1IJ50, 1IJ76, 1HB53, 1HD53, 1HZ53, 1HB55, 1HD55, 1HZ55, 1HZ85, 1HB54, 1HD54 |

<sup>a</sup> An enhanced version of ICD-10 developed by the Canadian Institute for Health Information (CIHI) for morbidity classification in Canada
<sup>b</sup> Canadian Classification of Health Interventions

**eExhibit 3.**
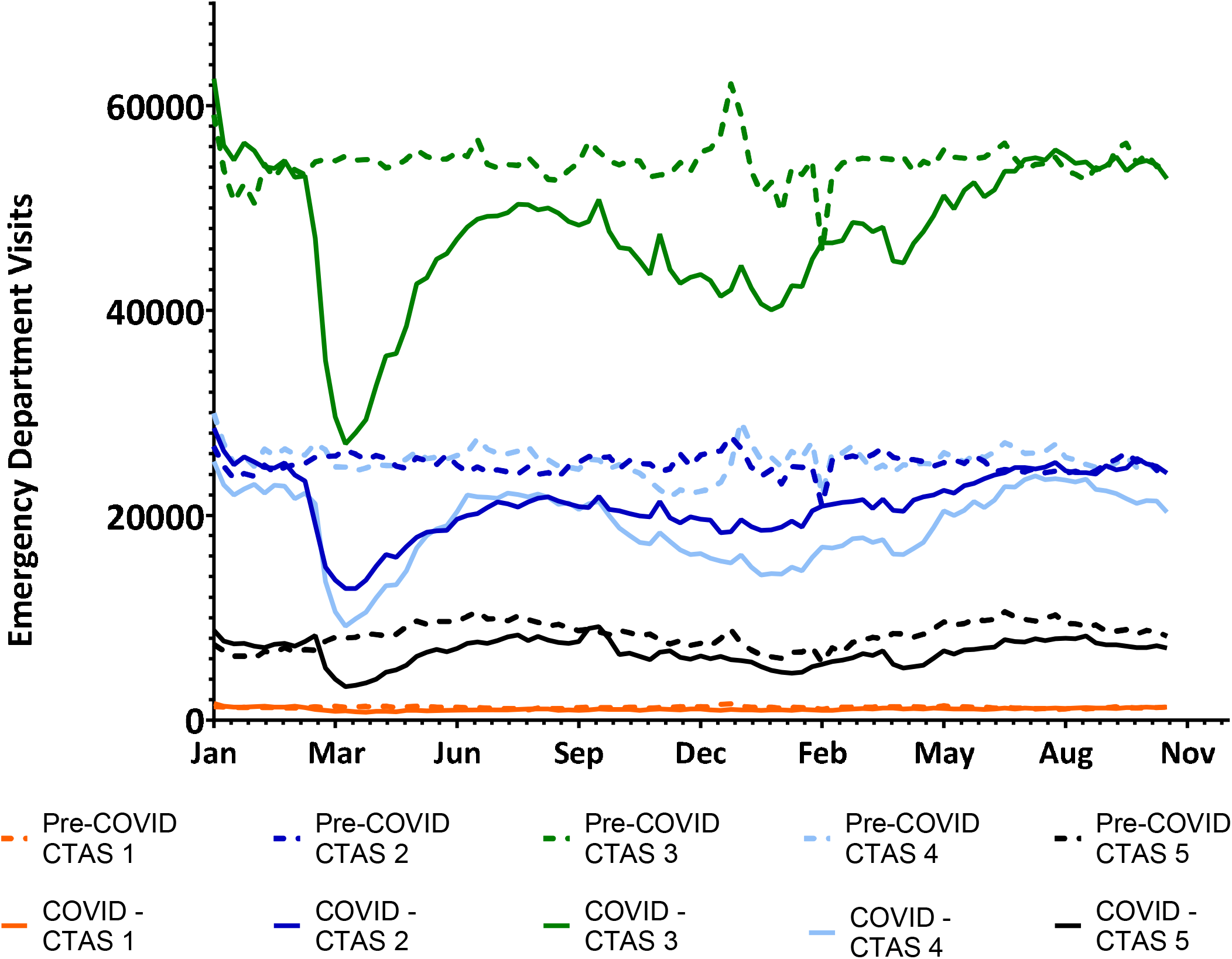
Weekly Emergency Department Visits by Canadian Triage and Acuity Scale (CTAS) Level, January 2019 to October 2021.

**eExhibit 4.**
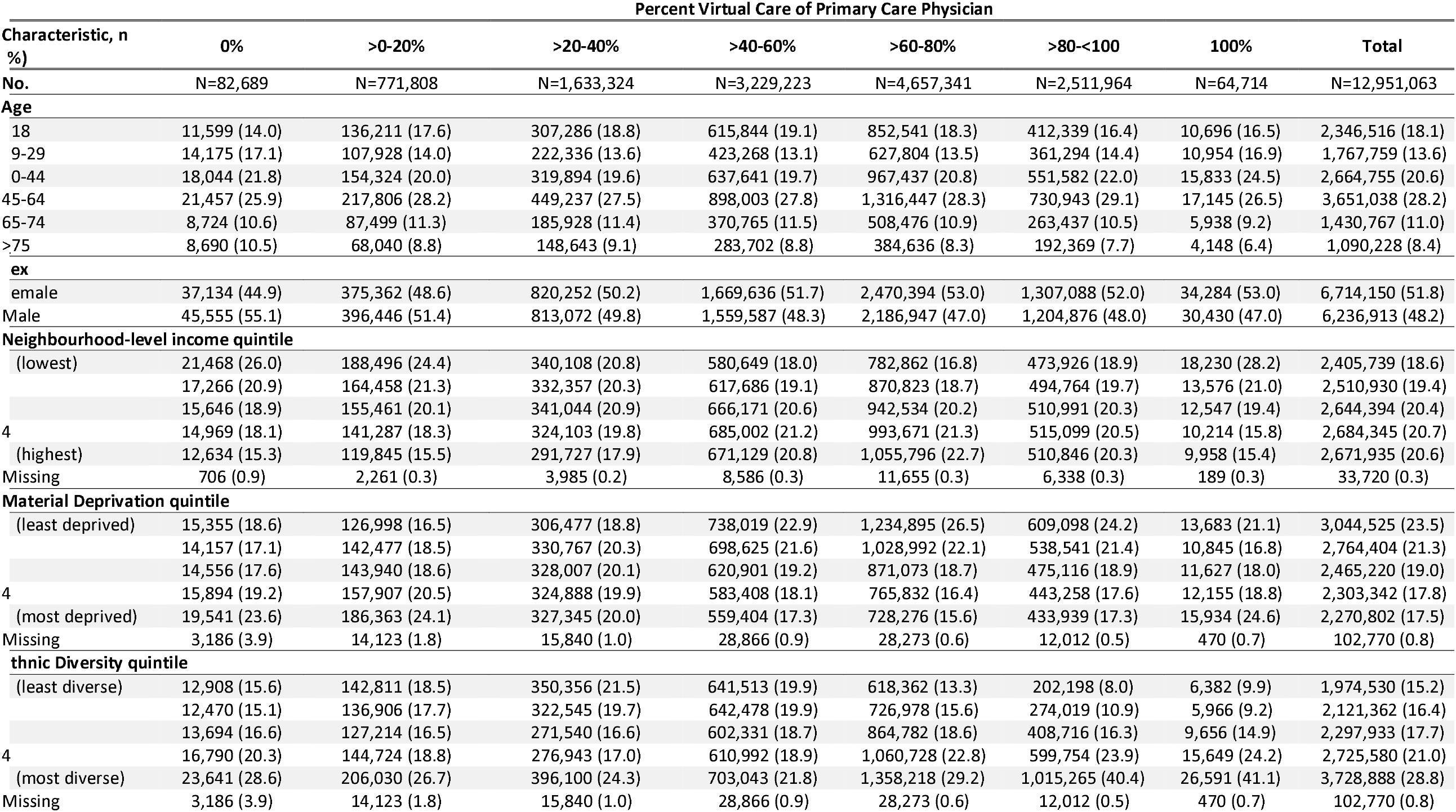
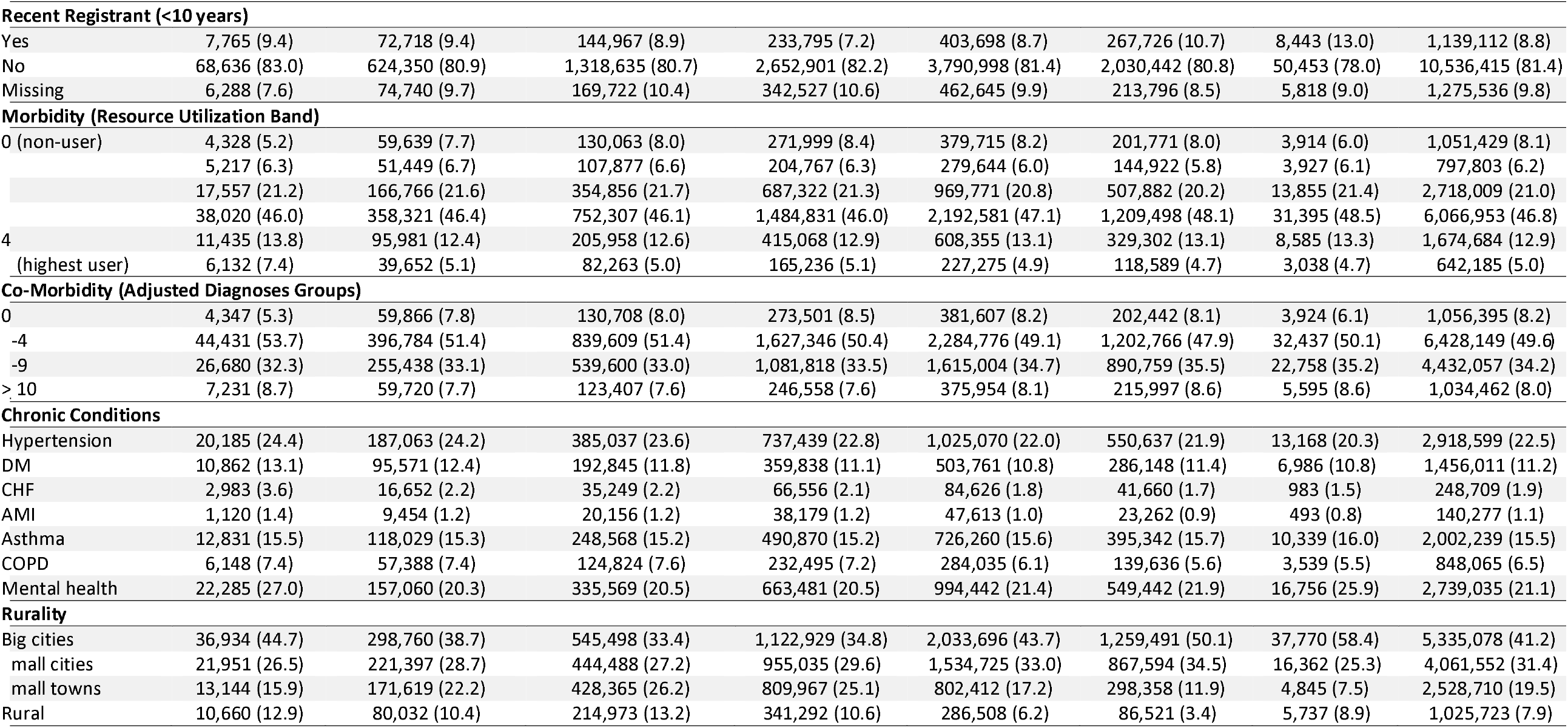
Patient Characteristics by the Percent of Physician Primary Care Provided Virtually, between February 1 and October 31, 2021, in Ontario, Canada.

**eExhibit 5a.** Physician and Practice Characteristics by the Percent of Care Provided Virtually, Comprehensive Family Physicians, between February 1 and October 31, 2021, in Ontario, Canada.

| Physician % Virtual Care | 0% | >0-20% | >20-40% | >40-60% | >60-80% | >80-<100% | 100% | Total |
| --- | --- | --- | --- | --- | --- | --- | --- | --- |
| **No. Physicians** | N=117 | N=544 | N=1,118 | N=2,265 | N=3,432 | N=1,881 | N=105 | N=9,462 |
| **No. Patients** | N=46,747 | N=661,761 | N=1,476,716 | N=2,966,294 | N=4,422,486 | N=2,386,349 | N=51,668 | N=12,125,959 |
| **Physician Age** |  |  |  |  |  |  |  |  |
| ≤44 | 33 (28.2) | 114 (21.0) | 378 (33.8) | 875 (38.6) | 1,319 (38.4) | 599 (31.8) | 43 (41.0) | 3,361 (35.5) |
| 45-64 | 34 (29.1) | 258 (47.4) | 512 (45.8) | 1,076 (47.5) | 1,674 (48.8) | 862 (45.8) | 30 (28.6) | 4,446 (47.0) |
| 65-74 | 37 (31.6) | 124 (22.8) | 179 (16.0) | 256 (11.3) | 369 (10.8) | 335 (17.8) | 23 (21.9) | 1,323 (14.0) |
| ≥75 | 13 (11.1) | 48 (8.8) | 49 (4.4) | 58 (2.6) | 70 (2.0) | 85 (4.5) | 9 (8.6) | 332 (3.5) |
| **Physician Sex** |  |  |  |  |  |  |  |  |
| Female | 31 (26.5) | 150 (27.6) | 438 (39.2) | 1,152 (50.9) | 1,953 (56.9) | 967 (51.4) | 68 (64.8) | 4,759 (50.3) |
| Male | 86 (73.5) | 394 (72.4) | 680 (60.8) | 1,113 (49.1) | 1,479 (43.1) | 914 (48.6) | 37 (35.2) | 4,703 (49.7) |
| **Primary Care Enrolment Model** |  |  |  |  |  |  |  |  |
| Non-team capitation | 8 (6.8) | 126 (23.2) | 317 (28.4) | 727 (32.1) | 1,190 (34.7) | 532 (28.3) | 11 (10.5) | 2,911 (30.8) |
| Enhanced fee-for-service | 19 (16.2) | 155 (28.5) | 297 (26.6) | 457 (20.2) | 876 (25.5) | 725 (38.5) | 23 (21.9) | 2,552 (27.0) |
| Team-based capitation | 12 (10.3) | 60 (11.0) | 286 (25.6) | 814 (35.9) | 1,044 (30.4) | 321 (17.1) | 9 (8.6) | 2,546 (26.9) |
| Traditional fee-for-service | 78 (66.7) | 203 (37.3) | 218 (19.5) | 267 (11.8) | 321 (9.4) | 303 (16.1) | 62 (59.0) | 1,452 (15.3) |
| Missing | 0 (0.0) | 0 (0.0) | 0 (0.0) | 0 (0.0) | 1 (0.0) | 0 (0.0) | 0 (0.0) | 1 (0.0) |
| **Rurality of Practice (RIO)** |  |  |  |  |  |  |  |  |
| Big cities | 54 (46.2) | 240 (44.1) | 430 (38.5) | 925 (40.8) | 1,797 (52.4) | 1,113 (59.2) | 66 (62.9) | 4,625 (48.9) |
| Small cities | 19 (16.2) | 146 (26.8) | 296 (26.5) | 643 (28.4) | 1,018 (29.7) | 558 (29.7) | 25 (23.8) | 2,705 (28.6) |
| Small towns | 25 (21.4) | 111 (20.4) | 250 (22.4) | 459 (20.3) | 403 (11.7) | 143 (7.6) | 7 (6.7) | 1,398 (14.8) |
| Rural | 19 (16.2) | 47 (8.6) | 142 (12.7) | 238 (10.5) | 214 (6.2) | 67 (3.6) | 7 (6.7) | 734 (7.8) |
| **Patient Panel Size** |  |  |  |  |  |  |  |  |
| <100 | 57 (48.7) | 61 (11.2) | 81 (7.2) | 122 (5.4) | 116 (3.4) | 89 (4.7) | 35 (33.3) | 561 (5.9) |
| 100-499 | 30 (25.6) | 103 (18.9) | 106 (9.5) | 169 (7.5) | 239 (7.0) | 200 (10.6) | 32 (30.5) | 879 (9.3) |
| 500-999 | 9 (7.7) | 85 (15.6) | 201 (18.0) | 490 (21.6) | 802 (23.4) | 443 (23.6) | 20 (19.0) | 2,050 (21.7) |
| 1000-1499 | 15 (12.8) | 104 (19.1) | 321 (28.7) | 654 (28.9) | 1,152 (33.6) | 520 (27.6) | 9 (8.6) | 2,775 (29.3) |
| 1500-1999 | - | 79 (14.5) | 212 (19.0) | 469 (20.7) | 675 (19.7) | 341 (18.1) | - | 1,785 (18.9) |
| ≥2000 | - | 112 (20.6) | 197 (17.6) | 361 (15.9) | 447 (13.0) | 288 (15.3) | - | 1,411 (14.9) |
| Missing | 0 (0.0) | 0 (0.0) | 0 (0.0) | 0 (0.0) | 1 (0.0) | 0 (0.0) | 0 (0.0) | 1 (0.0) |

**eExhibit 5b.**
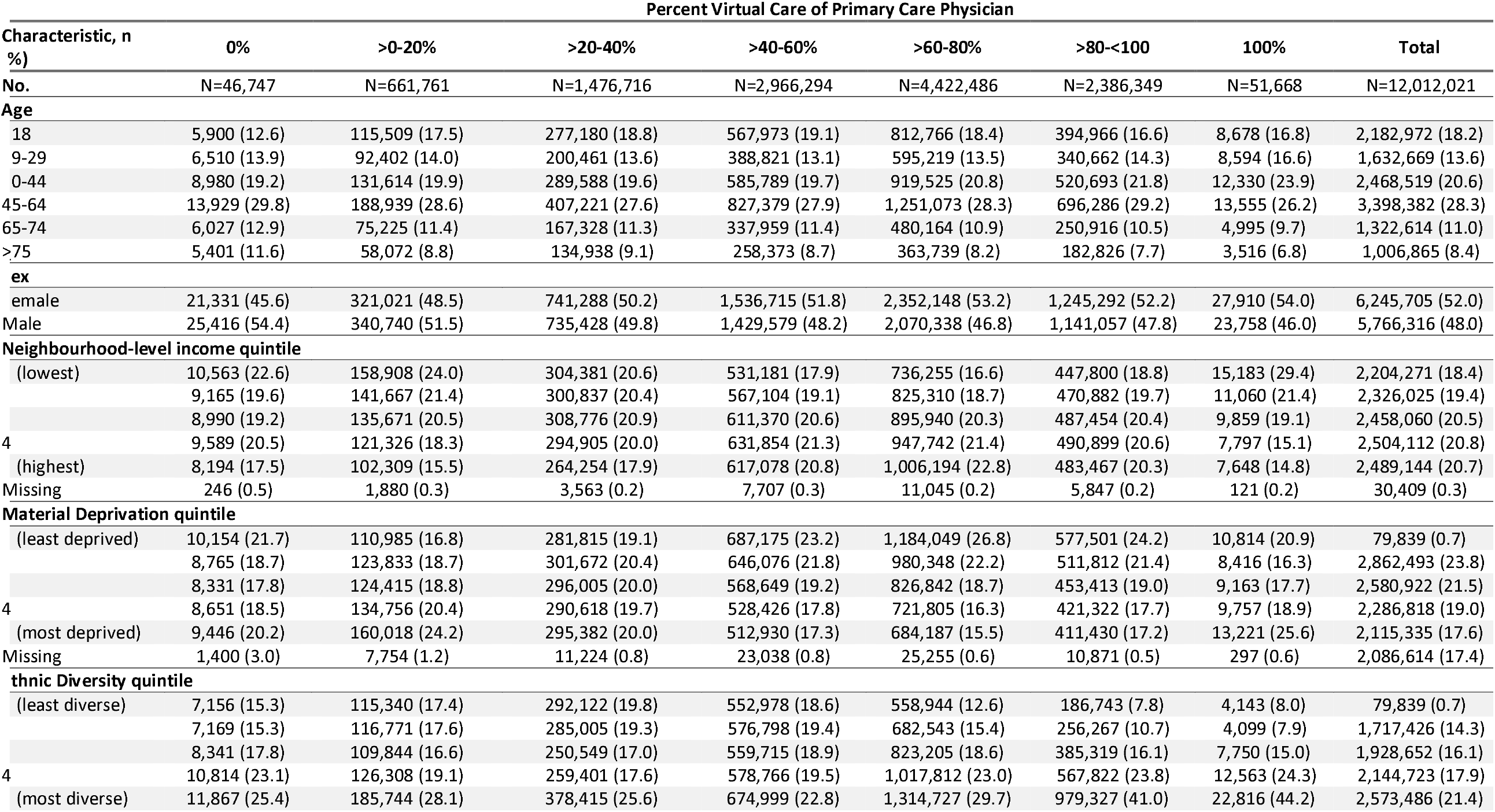
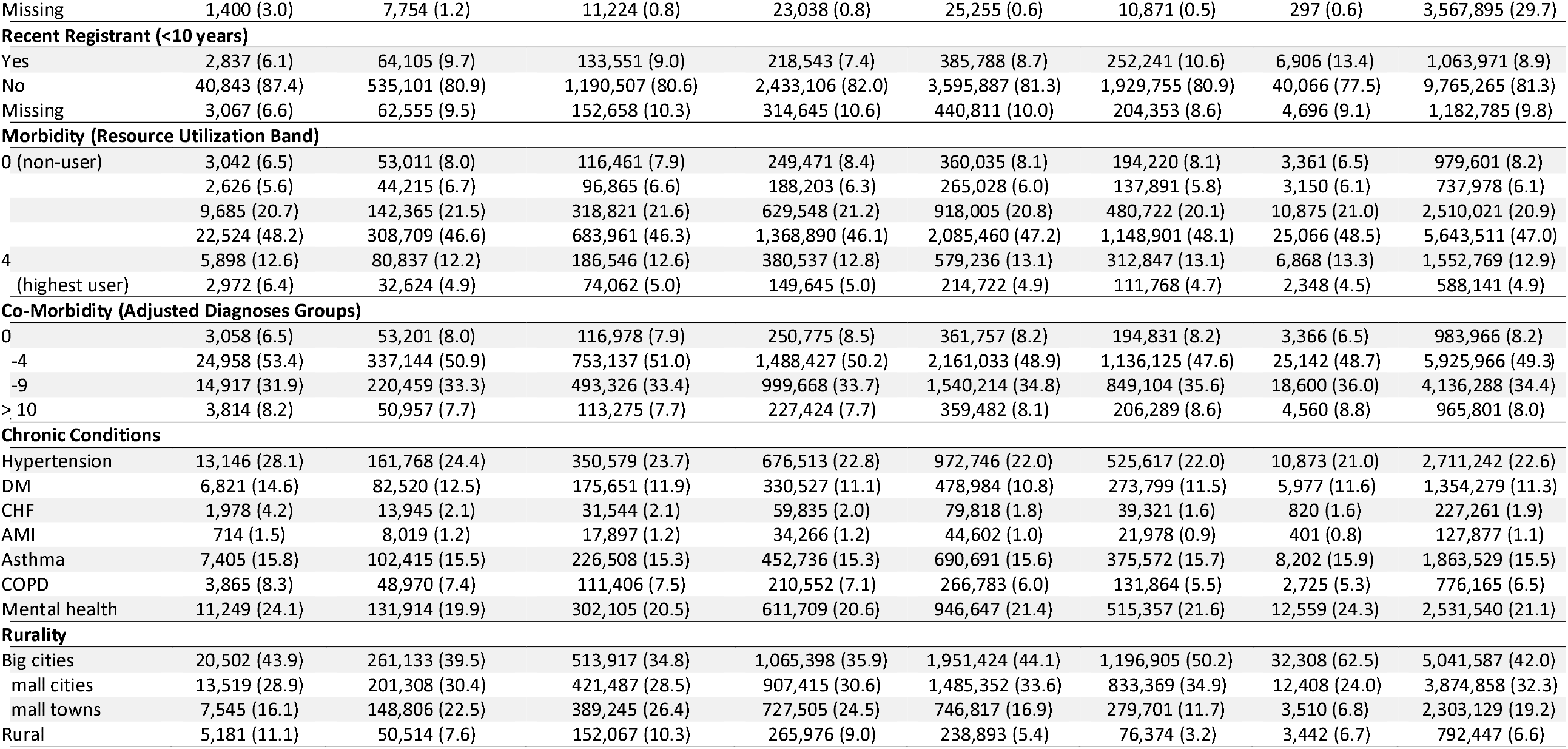
Characteristics of Patients Attached to Comprehensive Family Physicians by the Percent of Physician Primary Care Provided Virtually, between February 1 and October 31, 2021, in Ontario, Canada.

**eExhibit 6.** Health Service Utilization of Patients Attached to Comprehensive Family Physicians Stratified by the Attached Physician’s Percent of Care Provided Virtually, between February 1 and October 2021, in Ontario Canada.

| Physician % Virtual Care | 0% | >0-20% | >20-40% | >40-60% | >60-80% | >80-<100% | 100% | Total |
| --- | --- | --- | --- | --- | --- | --- | --- | --- |
| **No. Patients** | N=46,747 | N=661,761 | N=1,476,716 | N=2,966,294 | N=4,422,486 | N=2,386,349 | N=51,668 | N=12,125,959 |
| **Primary Care Services** |  |  |  |  |  |  |  |  |
| Any primary care visit, n (%) | 26,862 (57.5) | 411,991 (62.3) | 921,775 (62.4) | 1,858,118 (62.6) | 2,855,323 (64.6) | 1,569,605 (65.8) | 32,277 (62.5) | 7,675,951 (63.9) |
| Number of primary care visits per patient, mean ± SD | 2.3 ± 4.0 | 2.4 ± 3.9 | 2.4 ± 3.8 | 2.3 ± 3.6 | 2.6 ± 3.8 | 2.9 ± 4.2 | 2.8 ± 4.5 | 2.5 ± 3.9 |
| Percent of visits with usual family physician<sup>a</sup>, mean ± SD | 53.3 ± 45.2 | 67.7 ± 39.8 | 70.3 ± 38.1 | 71.7 ± 36.8 | 71.8 ± 36.6 | 68.2 ± 38.8 | 40.0 ± 43.0 | 70.4 ± 37.6 |
| **Other Health Care Use** |  |  |  |  |  |  |  |  |
| Any specialist visits, n (%) | 18,601 (39.8) | 249,150 (37.6) | 563,324 (38.1) | 1,150,948 (38.8) | 1,763,140 (39.9) | 959,637 (40.2) | 20,368 (39.4) | 4,725,168 (39.3) |
| Ambulatory care-sensitive condition visits, n (%) | 164 (0.4) | 1,734 (0.3) | 3,675 (0.2) | 6,968 (0.2) | 9,030 (0.2) | 4,393 (0.2) | 91 (0.2) | 26,055 (0.2) |
| **Emergency Department Utilization, February to October 2019/2021** |  |  |  |  |  |  |  |  |
| Total visits per 1,000, 2021, mean ± SD<sup>b</sup> | 348.0 ± 1204.0 | 287.2 ± 981.7 | 282.4 ± 1005.9 | 274.7 ± 893.5 | 248.9 ± 810.0 | 238.0 ± 780.5 | 262.1 ± 972.0 | 259.8 ± 864.4 |
| Total visits per 1,000, 2019, mean ± SD<sup>b</sup> | 387.9 ± 1293.1 | 335.5 ± 1023.4 | 330.3 ± 959.2 | 317.4 ± 945.7 | 284.1 ± 911.7 | 268.7 ± 828.6 | 282.2 ± 853.4 | 298.4 ± 920.0 |
| Absolute difference 2021 vs 2019 (95% CI) | -39.9 (-54.3, -25.6) | -48.3 (-51.8, -44.9) | -47.9 (-50.2, -45.6) | -42.7 (-44.2, -41.2) | -35.2 (-36.4, -34.0) | -30.8 (-32.3, -29.3) | -20.0 (-30.2, -9.9) | -38.6 (-39.3, -37.9) |
| Percent change 2021 vs 2019 (95% CI) | -10.3 (-13.8, -6.8) | -14.4 (-15.4, -13.4) | -14.5 (-15.2, -13.9) | -13.5 (-13.9, -13.0) | -12.4 (-12.8, -12.0) | -11.5 (-12.0, -10.9) | -7.1 (-10.7, -3.5) | -12.9 (-13.2, -12.7) |

<sup>a</sup> Among patients with 2 or more visits between February and October 2021
<sup>b</sup> This includes only patients whose attached physicians had claims in both 2021 and 2019.
Abbreviations: SD, standard deviation; CI, confidence interval

**eExhibit 7.**
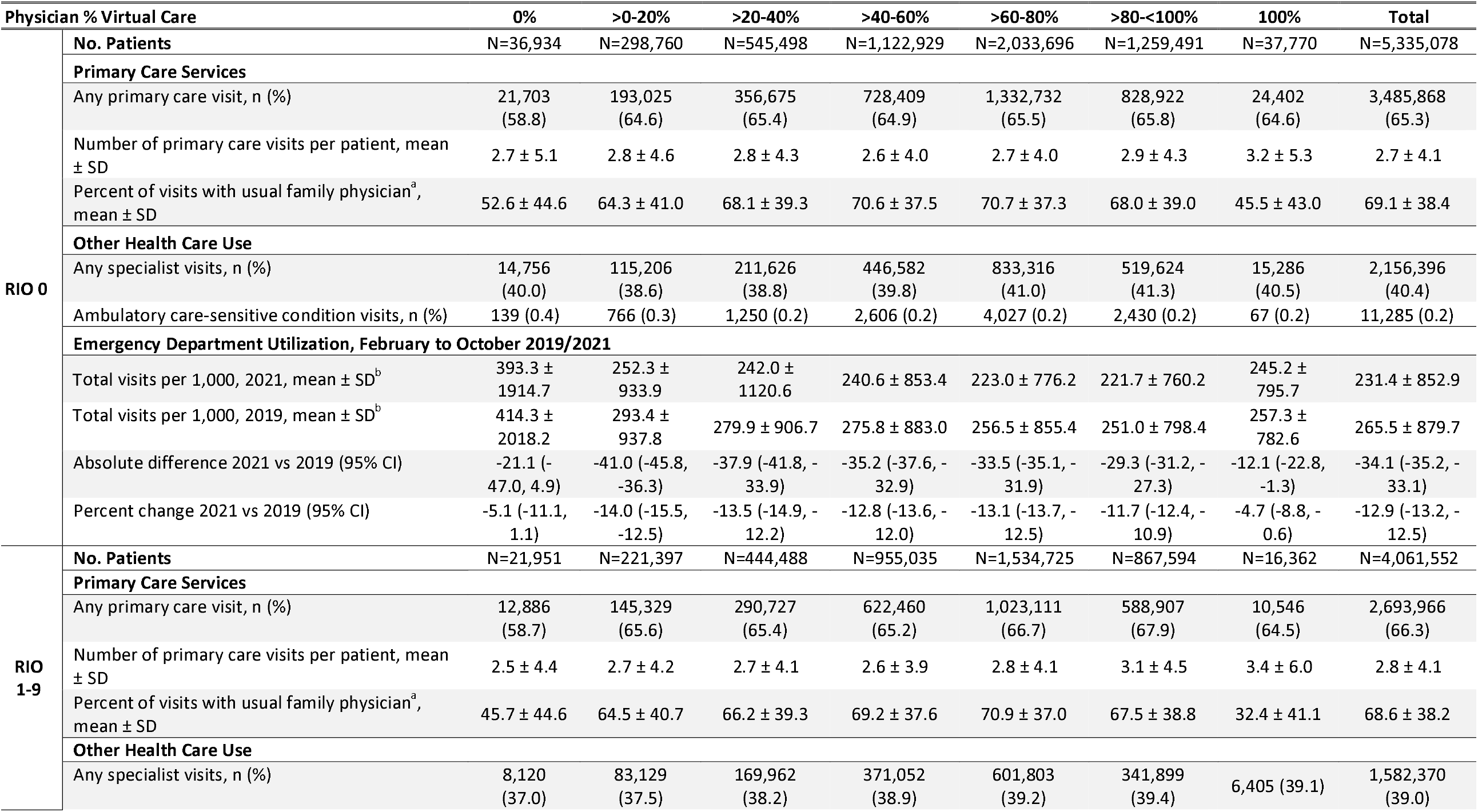
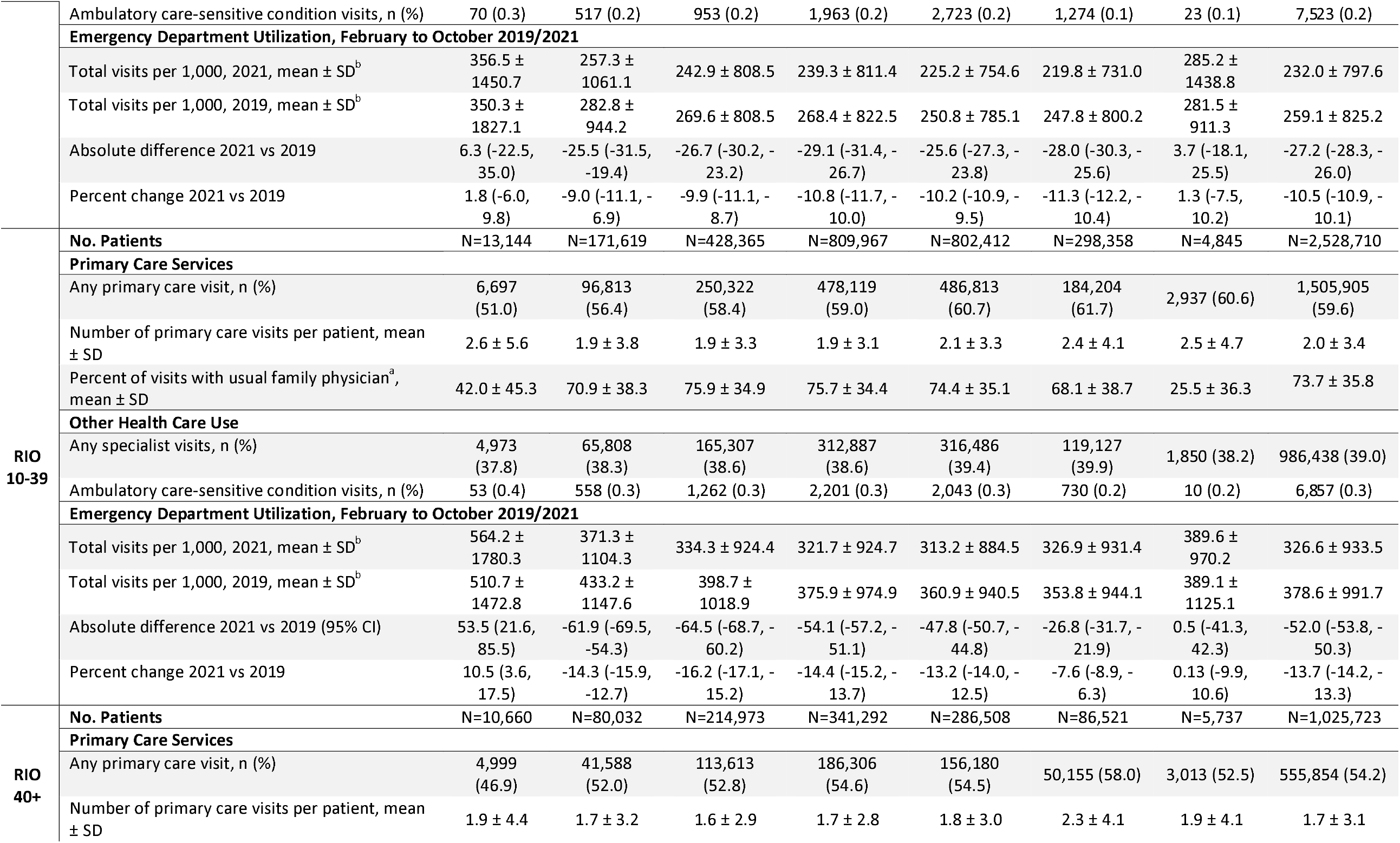
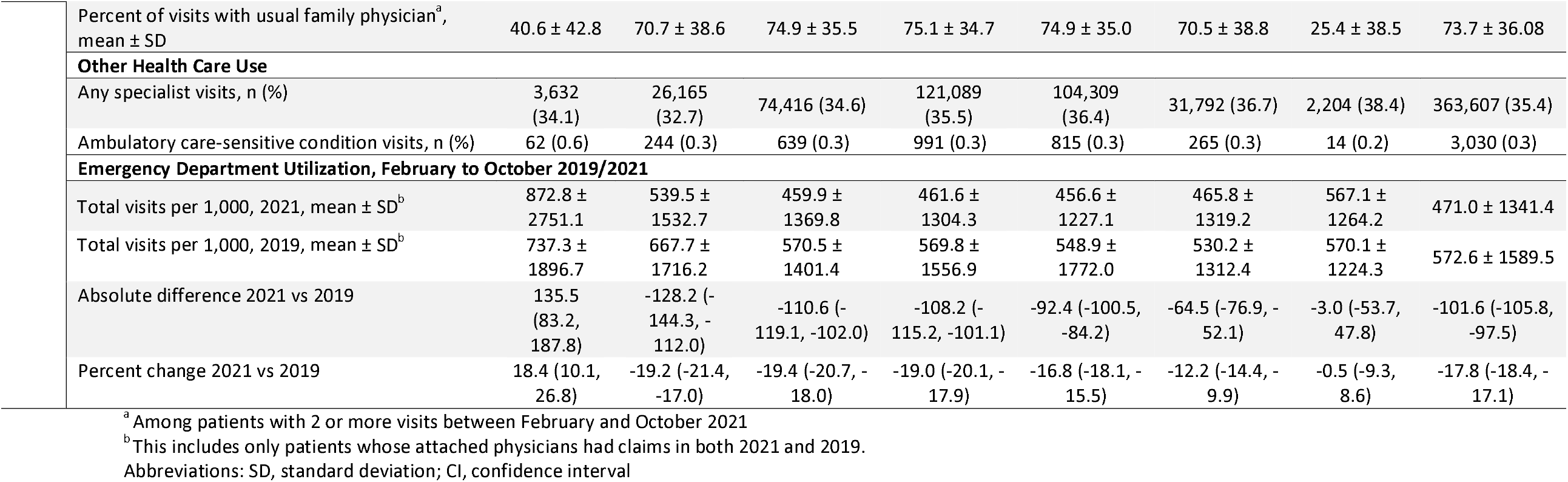
Patient Health Service Utilization Stratified by the Attached Physician’s Percent of Care Provided Virtually, between February 1 and October 2021, in Ontario Canada, by Rurality.

STROBE Statement—Checklist of items that should be included in reports of ***cross-sectional studies***

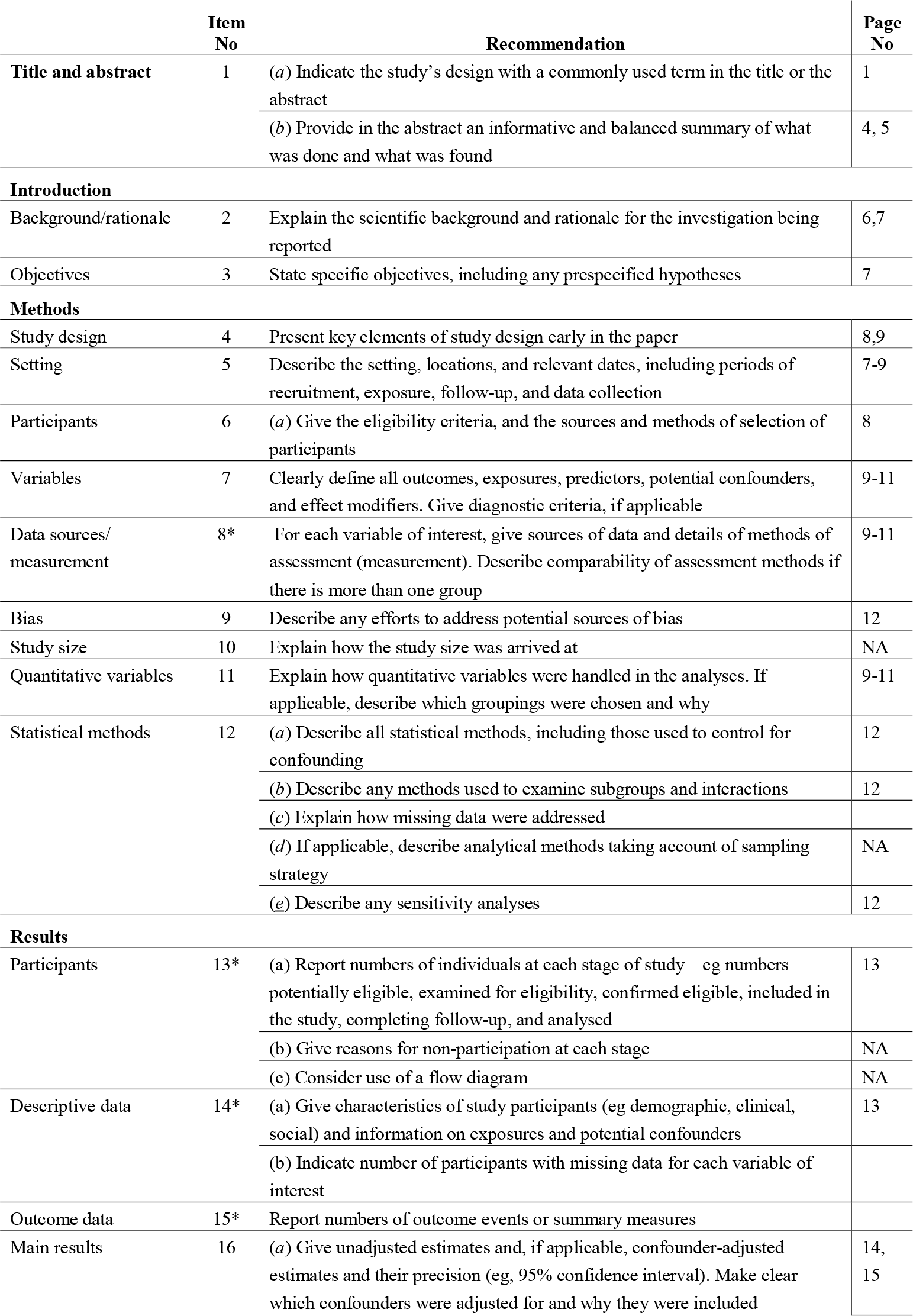
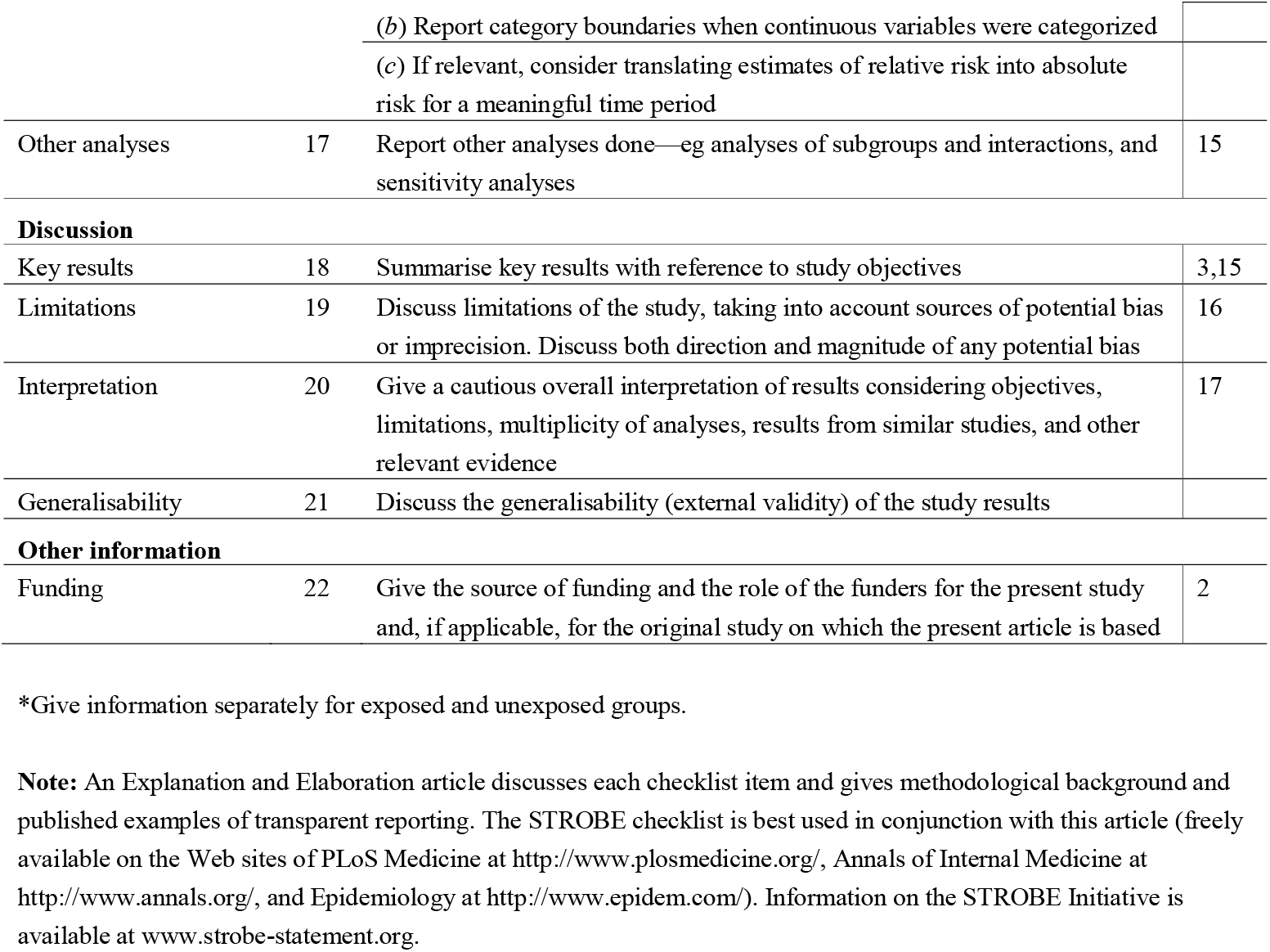

## References

1. Glazier RH, Green ME, Wu FC, Frymire E, Kopp A, Kiran T. Shifts in office and virtual primary care during the early COVID-19 pandemic in Ontario, Canada. CMAJ. 2021;193(6):E200–E210.

2. Canadians’ Health Care Experiences During COVID-19. Canada Health Infoway. https://www.infoway-inforoute.ca/en/component/edocman/resources/reports/benefits-evaluation/3828-canadians-health-care-experiences-during-covid-19. Published 2022. Accessed July, 2022.

3. Agarwal P, Wang R, Meaney C, et al. Sociodemographic differences in patient experience with primary care during COVID-19: results from a cross-sectional survey in Ontario, Canada. BMJ Open. 2022;12(5):e056868.

4. Nguyen M, Waller M, Pandya A, Portnoy J. A Review of Patient and Provider Satisfaction with Telemedicine. Curr Allergy Asthma Rep. 2020;20(11):72.

5. Eze ND, Mateus C, Cravo Oliveira Hashiguchi T. Telemedicine in the OECD: An umbrella review of clinical and cost-effectiveness, patient experience and implementation. PLoS One. 2020;15(8):e0237585.

6. Guinart D, Marcy P, Hauser M, Dwyer M, Kane JM. Patient Attitudes Toward Telepsychiatry During the COVID-19 Pandemic: A Nationwide, Multisite Survey. JMIR Ment Health. 2020;7(12):e24761.

7. Khoong EC, Sharma AE, Gupta K, Adler-Milstein J, Sarkar U. The Abrupt Expansion of Ambulatory Telemedicine: Implications for Patient Safety. J Gen Intern Med. 2022;37(5):1270–1274.

8. Eberly LA, Kallan MJ, Julien HM, et al. Patient Characteristics Associated With Telemedicine Access for Primary and Specialty Ambulatory Care During the COVID-19 Pandemic. JAMA Netw Open. 2020;3(12):e2031640.

9. Zhang D, Shi L, Han X, et al. Disparities in telehealth utilization during the COVID-19 pandemic: Findings from a nationally representative survey in the United States. J Telemed Telecare. 2021:1357633×211051677.

10. Carr MJ, Wright AK, Leelarathna L, et al. Impact of COVID-19 restrictions on diabetes health checks and prescribing for people with type 2 diabetes: a UK-wide cohort study involving 618 161 people in primary care. BMJ Qual Saf. 2022;31(7):503–514.

11. Li KY, Ng S, Zhu Z, McCullough JS, Kocher KE, Ellimoottil C. Association Between Primary Care Practice Telehealth Use and Acute Care Visits for Ambulatory Care-Sensitive Conditions During COVID-19. JAMA Netw Open. 2022;5(3):e225484.

12. Dupraz J, Le Pogam MA, Peytremann-Bridevaux I. Early impact of the COVID-19 pandemic on in-person outpatient care utilisation: a rapid review. BMJ Open. 2022;12(3):e056086.

13. Chu C, Cram P, Pang A, Stamenova V, Tadrous M, Bhatia RS. Rural Telemedicine Use Before and During the COVID-19 Pandemic: Repeated Cross-sectional Study. J Med Internet Res. 2021;23(4):e26960.

14. Almathami HKY, Win KT, Vlahu-Gjorgievska E. Barriers and Facilitators That Influence Telemedicine-Based, Real-Time, Online Consultation at Patients’ Homes: Systematic Literature Review. J Med Internet Res. 2020;22(2):e16407.

15. Adepoju OE, Tran L, Agwuncha R, et al. Associations between Patient- and Provider Level Factors, and Telemedicine Use in Family Medicine Clinics. J Am Board Fam Med. 2022;35(3):457–464.

16. Alami H, Gagnon MP, Wootton R, Fortin JP, Zanaboni P. Exploring factors associated with the uneven utilization of telemedicine in Norway: a mixed methods study. BMC Med Inform Decis Mak. 2017;17(1):180.

17. Liu L, Goodarzi Z, Jones A, Posno R, Straus SE, Watt JA. Factors associated with virtual care access in older adults: a cross-sectional study. Age Ageing. 2021;50(4):1412–1415.

18. COVID-19 UPDATE REGARDING THE PROVISION OF IN-PERSON AND VIRTUAL CARE. CPSO. https://www.cpso.on.ca/en/News/COVID-19-Updates/Information-for-Physicians/COVID-19-update-regarding-the-provision-of-in-(1). Published 2021. Accessed June, 2022.

19. Population estimates on July 1st, by age and sex1, 2, 3, 4. Statistics Canada. https://www150.statcan.gc.ca/t1/tbl1/en/tv.action?pid=1710000501&pickMembers%5B0%5D=1.7&pickMembers%5B1%5D=2.1. Published 2021. Accessed June, 2022.

20. Kiran T, Kopp A, Moineddin R, Glazier RH. Longitudinal evaluation of physician payment reform and team-based care for chronic disease management and prevention. Cmaj. 2015;187(17):E494–e502.

21. Population distribution of Ontario in 2016, by rural/urban type. Statista Research Department. https://www.statista.com/statistics/608698/population-distribution-of-ontario-by-rural-urban-type/. Published 2022. Accessed July, 2022.

22. Kiran T, Moineddin R, Kopp A, Glazier RH. Impact of Team-Based Care on Emergency Department Use. Ann Fam Med. 2022;20(1):24–31.

23. Ontario COVID-19 Data Tool. Public Health Ontario. https://www.publichealthontario.ca/en/data-and-analysis/infectious-disease/covid-19-data-surveillance/covid-19-data-tool?tab=trends. Published 2022. Accessed July, 2022.

24. Kralj B. Measuring rurality - RIO 2008_BASIC : methodology and results. Toronto, Ont.: Ontario Medical Association Economics Department; 2013.

25. Matheson FI, Dunn JR, Smith KL, Moineddin R, Glazier RH. Development of the Canadian Marginalization Index: a new tool for the study of inequality. Can J Public Health. 2012;103(8 Suppl 2):S12–16.

26. Ontario Marginalization Index (ON-Marg). Ontario Community Health Profiles Partnership. https://www.ontariohealthprofiles.ca/onmargON.php. Accessed July, 2022.

27. Johns Hopkins ACG® System. Baltimore: Johns Hopkins University. https://www.hopkinsacg.org/. Accessed July, 2022.

28. Term: Resource Utilization Bands (RUBs). University of Manitoba. http://mchp-appserv.cpe.umanitoba.ca/viewDefinition.php?definitionID=104613. Published 2015. Accessed July, 2022.

29. Schultz SE, Glazier RH. Identification of physicians providing comprehensive primary care in Ontario: a retrospective analysis using linked administrative data. CMAJ Open. 2017;5(4):E856–e863.

30. Dayal P, Chang CH, Benko WS, et al. Hospital Utilization Among Rural Children Served by Pediatric Neurology Telemedicine Clinics. JAMA Netw Open. 2019;2(8):e199364.

31. Shah MN, Wasserman EB, Gillespie SM, et al. High-Intensity Telemedicine Decreases Emergency Department Use for Ambulatory Care Sensitive Conditions by Older Adult Senior Living Community Residents. J Am Med Dir Assoc. 2015;16(12):1077–1081.

32. Friedman RSC, Carpenter DM, Shaver JM, McDermott SC, Voelkel J. Telemedicine Familiarity and Post-Disaster Utilization of Emergency and Hospital Services for Ambulatory Care Sensitive Conditions. Am J Prev Med. 2022;63(1):e1–e9.

33. Stamenova V, Chu C, Pang A, et al. Virtual care use during the COVID-19 pandemic and its impact on healthcare utilization in patients with chronic disease: A population-based repeated cross-sectional study. PLoS One. 2022;17(4):e0267218.

34. Crawley M. Family doctors deny they’re causing Ontario’s emergency room problems. CBC NEWS. https://www.cbc.ca/news/canada/toronto/ontario-hospital-emergency-room-family-doctors-1.6516769. Published 2022. Accessed July, 2022.

